# Integrative effects of Telomere Length, Epigenetic Age, and Mitochondrial DNA abundance in Alzheimer’s Disease

**DOI:** 10.1101/2025.07.16.25331683

**Authors:** Shea J Andrews, Rakshya U Sharma, Brendan A Mitchell, Tong Tong, Luke W. Bonham, Alan E Renton, Xiaoling Zhang, Marina Sirota, Duygu Tosun, Kristine Yaffe, the Alzheimer’s Disease Neuroimaging Initiative

**Author notes:** Corresponding author Shea Andrews, PhD Assistant Professor, Department of Psychiatry and Behavioral Sciences University of California, San Francisco, Nancy Friend Pritzker Psychiatry Building 675 18th St.

## Abstract

**Background and Objectives:** Biological age, reflecting the cumulative molecular and cellular damage such as telomere attrition, epigenetic alterations and mitochondrial dysfunction, may better capture ageLrelated decline and Alzheimer’s disease (AD) risk than chronological age. Most studies have focused on one measure of biological age and not investigated joint or interactive contributions to AD pathogenesis.

**Methods:** We estimated bloodLderived telomere length (TL) via qPCR, epigenetic age (DNAm age) using the CausAge clock, and mitochondrial DNA copy number (mtDNAcn) from whole genome sequencing in 640 participants from the Alzheimer’s Disease Neuroimaging Initiative (ADNI; Age: 74.91±7.56, Female: 44.8%, Cognitively Unimpaired: 34.3%, Mild Cognitive Impairment: 52%, AD: 12.9%). Linear mixedLeffects models examined the associations and interactions of these markers with cognitive decline for memory, executive function, language ability, visuospatial ability, and global cognition, while linear regression tested associations with cross-sectional AD biomarkers (CSF Aβ_42_, totalLtau, pTau_181_, and meta-ROI for cortical thickness and gray matter volume). Models adjusted for baseline age, sex, clinical dementia rating scale, *APOE*, blood cell composition, and outcomeLspecific covariates (education and intracranial volume).

**Results:** Individually, TL and DNAm age, were not associated with cognition, CSF biomarkers, or neuroimaging outcomes, while higher mtDNAcn was associated with lower CSF tau and pTau_181_. Interaction models revealed that mtDNAcn modified the effects of both TL and DNAm age: at higher mtDNAcn, shorter TL predicted poorer global cognition (β = 0.033 ± 0.014, p = 0.020) and older DNAm age predicted poorer language performance (β = –0.059 ± 0.028, p = 0.038). A significant three-way interaction showed that the combination of higher mtDNAcn, longer TL, and older DNAm age was associated with lower grey-matter volume.

**Discussion:** These findings suggest that increased mtDNAcn may act as a compensatory response to accelerated epigenetic aging and telomere attrition. Our results underscore the importance of evaluating the interplay among multiple biological aging markers when investigating AD pathogenesis.

**Glossary:** mtDNAcn: mitochondrial DNA copy number; TL: telomere length; AD: Alzheimer’s disease; CDR-SB: Clinical dementia rating sum of boxes; CSF: cerebrospinal fluid

## Introduction

Age is one of the strongest risk factors for Alzheimer’s disease and Alzheimer’s disease-related dementias (AD/ADRD), with dementia incidence increasing sharply with advancing chronological age ^1^. The aging process is driven by the accumulation of molecular and cellular damage, which proceeds at different rates among individuals and defines a person’s biological age ^2^. The “hallmarks of aging” comprise 12 independent biological processes characterized by such damage, including telomere attrition, epigenetic alterations, and mitochondrial dysfunction, which have been associated with an increased risk of dementia ^3^. However, how multiple measures and their interactions among these aging hallmarks collectively influence AD pathogenesis remains poorly understood.

DNA methylation (DNAm) is a heritable yet dynamic epigenetic modification that modulates gene expression, primarily through the addition of methyl groups to the 5-position cytosine residues within CpG dinucleotides ^3^. Epigenetic clocks estimate biological age, termed epigenetic age (DNAm age), by quantifying age-associated changes in DNAm patterns at specific CpG sites ^4^. Accelerated epigenetic aging (i.e., the difference between DNAm age and chronological age) has been associated with increased dementia risk ^5^, poorer cognitive performance ^5^, reduced total brain and hippocampal volume ^6,7^, and AD neuropathology ^8^. Telomeres are nucleoprotein structures capping the ends of chromosomes that are essential for maintaining genomic integrity ^9^. Telomeres progressively shorten with successive cell divisions, and shorter telomere length (TL) has been associated with increased AD/ADRD risk, lower cognitive performance, smaller hippocampal volume, and reduced total brain tissue volume ^10–13^. Mitochondrial dysfunction is an early and prominent feature of AD characterized by impaired glucose metabolism, dysregulated mitochondrial fusion/fission, elevated oxidative stress, disturbed Ca2+ homeostasis, and altered expression of nuclear-encoded mitochondrial genes and proteins ^14^. Mitochondrial DNA copy number (mtDNAcn) – the number of mitochondrial genomes per cell – serves as a biomarker of mitochondrial activity and a proxy for overall mitochondrial function ^15^. Lower blood-based mtDNAcn at baseline is associated with increased risk of dementia 17 years later ^16^ and worse cognitive performance ^17^, while lower brain-derived mtDNAcn was associated with higher odds of AD neuropathology ^14,18^.

While TL, mtDNAcn, and epigenetic age are individually associated with AD pathogenesis, these hallmarks of aging are mechanistically interconnected. Mitochondria produce reactive oxygen species (ROS) that can induce DNA damage, thereby accelerating telomere shortening^19^. Shortened telomeres can activate the tumor suppressor gene p53, which subsequently inhibits PGC-1α—a master regulator of mitochondrial biogenesis—leading to decreased mitochondrial function, reduced ATP production, and further elevation of ROS levels ^20,21^. Furthermore, age-related epigenetic changes can alter the expression of nuclear-encoded mitochondrial genes, impacting mitochondrial function ^22^. Conversely, metabolic signals originating from mitochondria can trigger epigenetic modifications within the nuclear genome ^23,24^. Accelerated epigenetic aging is associated with telomere attrition ^25,26^, and individuals with telomere biology disorders often exhibit accelerated epigenetic age, suggesting premature cellular aging ^27^. Thus, a comprehensive understanding of biological aging’s impact on AD pathogenesis necessitates evaluating the interplay among these key aging hallmarks.

Using the Alzheimer’s Disease Neuroimaging Initiative (ADNI) cohort, we investigated whether TL, DNAm age, and mtDNAcn interact to influence AD endophenotypes—cognitive performance, core cerebrospinal-fluid biomarkers, and MRI-based neurodegeneration measures—over and above the independent contribution of each biomarker.

## Methods

### Alzheimer’s Disease Neuroimaging Initiative

Genomic and clinical data were obtained from the ADNI, a longitudinal, observational study launched in 2004 as a public-private partnership, enrolling participants at over 50 sites across the United States and Canada and has been described in detail previously ^28^.

This study leveraged data from ADNI participants for whom whole-genome sequencing, DNAm profiling, telomere length measurements, cognitive assessments, CSF biomarker acquisition, and magnetic resonance imaging data were available. TL and DNAm were generated in a subset of participants with longitudinal DNA samples, enriched for diagnostic conversion ^29,30^. WGS was conducted in phased efforts and was available for a subset across diagnostic groups without explicit conversion enrichment ^28^. Core clinical, cognitive, and MRI assessments were collected longitudinally in the main ADNI cohort, whereas CSF collection was optional and phase-dependent ^28,31–33^. For the present study,‘baseline’ was defined as the time point corresponding to the isolation of DNA from which genomic data were generated. Up to 640 participants with the required genomic and clinical data were included, including 216 cognitively unimpaired older adults (CU), 330 individuals with mild cognitive impairment (MCI), and 94 individuals with a clinical diagnosis of dementia due to AD (Table 1). Clinical symptoms severity was assessed using the Clinical Dementia Rating Scale Sum of Boxes (CDR-SB) score.

**Table 1:** Demographic, clinical, and biomarker characteristics of the study cohort.

| Variable | N | CU<br>N = 216 | MCI<br>N= 330 | Dementia<br>N = 94 | p-value<sup>†</sup> |
| --- | --- | --- | --- | --- | --- |
| **Demographics** |  |  |  |  |  |
| Visits* | 640 | 6 (2) | 6 (2) | 3 (1) | <0.001 |
| Age | 640 | 77 (7) | 73 (8) | 77 (8) | <0.001 |
| Female | 640 | 108 (50%) | 143 (43%) | 32 (34%) | 0.031 |
| Education | 640 |  |  |  | 0.6 |
| High School |  | 23 (11%) | 47 (14%) | 15 (16%) |  |
| College |  | 100 (46%) | 136 (41%) | 39 (41%) |  |
| Graduate |  | 93 (43%) | 147 (45%) | 40 (43%) |  |
| *APOE* | 640 |  |  |  | <0.001 |
| ε3/ε3 |  | 136 (63%) | 151 (46%) | 29 (31%) |  |
| ε2+ |  | 27 (13%) | 27 (8.2%) | 1 (1.1%) |  |
| ε4+ |  | 53 (25%) | 152 (46%) | 64 (68%) |  |
| Memory | 640 | 0.95 (0.52) | 0.33 (0.61) | -0.92 (0.60) | <0.001 |
| Executive Function | 640 | 0.72 (0.46) | 0.41 (0.57) | -0.56 (0.77) | <0.001 |
| Language | 640 | 0.82 (0.49) | 0.44 (0.51) | -0.29 (0.73) | <0.001 |
| Visual Spatial | 640 | 0.59 (0.45) | 0.44 (0.53) | -0.12 (0.76) | <0.001 |
| **MRI** |  |  |  |  |  |
| Cortical Thickness, mm | 588 | 2.51 (0.14) | 2.46 (0.19) | 2.21 (0.19) | <0.001 |
| Gray Matter Volume, mm<sup>3</sup> | 588 | 28,054 (3,470) | 27,279 (4,486) | 22,333 (3,803) | <0.001 |
| **CSF<sup>‡</sup>** |  |  |  |  |  |
| Aβ<sub>42</sub> | 448 | 0.43 (0.92) | -0.07 (0.95) | -0.59 (0.82) | <0.001 |
| Tau | 433 | -0.45 (0.83) | -0.09 (1.05) | 0.80 (0.81) | <0.001 |
| pTau<sub>181</sub> | 448 | -0.09 (0.90) | 0.25 (1.10) | 0.68 (0.87) | <0.001 |
| **Biological Aging** |  |  |  |  |  |
| Telomere Length | 640 | 5,354 (417) | 5,405 (403) | 5,384 (393) | 0.2 |
| mtDNAcn | 624 | 161 (35) | 169 (52) | 152 (29) | 0.009 |
| CausAge | 640 | 63.5 (5.7) | 61.7 (5.8) | 64.6 (7.3) | <0.001 |

<sup>†</sup>Comparisons across diagnostic groups using Kruskal-Wallis rank sum test, Pearson's Chi-squared test, or Fisher's exact test. \*Average number of visits from DNA isolation. <sup>‡</sup> z-normalized pg/mL

### Standard Protocol Approvals, Registrations, and Patient Consents

The ADNI study was approved by the ethical standard committee at each participating site, and all participants provided written informed consent for imaging and genetic sample collection, and questionnaires, as approved by each site’s institutional review board. ADNI investigators contributed to the study design and data collection but were not involved in the analysis or writing of this report.

### AD Endophenotypes

#### Cognitive Performance

Participants undergo an extensive neuropsychological battery at visit including the Mini-Mental State Examination (MMSE), Alzheimer’s Disease Assessment Schedule–Cognition (ADAS-Cog), Boston Naming Test, Rey Auditory Verbal Learning Test (RAVLT), Wechsler Memory Scale–Revised (WMS-R) Digit Span, WMS-R Logical Memory, Trails A & B, clock drawing, animal fluency, and the Montreal Cognitive Assessment (MoCA) ^28,31^. Using this test battery, the Alzheimer’s Diseases Sequencing Project - Phenotype Harmonization Consortium (ADSP-PHC) has generated harmonized cognitive domain scores ^34^. Briefly, a panel of content experts considers every test from each protocol and assigns it to a single primary cognitive domain – either memory, executive functioning, language, visuospatial ability, or other. Tests administered consistently across cohorts serve as anchor items and facilitate co-calibration. Confirmatory factor analysis is applied to each cognitive domain, choosing between models based on fit statistics and person-level impact, producing co-calibrated scores for memory, executive functioning, language, and visuospatial functioning. We further created a global cognitive domain score by averaging the domain scores.

#### CSF Amyloid and tau biomarkers

CSF was collected by lumbar puncture and levels of Ab_42_, total-tau (tau), and phosphorylated tau (pTau_181_) were quantified on the multiplex xMAP Luminex platform (Luminex Corp, Austin, TX) and Innogenetics INNO-BIA Alz-Bio3 (Innogenetics, Ghent, Belgium) immunoassay reagents, as described previously ^32^. Harmonization of the raw CSF levels was performed by the ADSP-PHC by log-transforming the protein values, removing outliers (defined as values below Q1 − 1.5×IQR or above Q3 + 1.5×IQR, where Q1 and Q3 are the first and third quartiles, respectively), and then z-standardizing the protein values ^35^. The current analyses used the closest CSF collection to the baseline genomics measurements; every included CSF measurement was obtained within one year of DNA isolation.

#### Neuroimaging

The ADSP-PHC conducted pre-processing and quality control of T1 MRI scans via Freesurfer, with longitudinal COMBAT used to harmonize across scanner platforms and models, site, and field strengths ^36^. We constructed an AD-Signature composite that combines measures of cortical thickness and gray matter volume across six regions ^33^. For gray matter volume, we averaged the right and left volumes for the hippocampus, amygdala, entorhinal, middle temporal, inferior temporal, and temporal pole; and then summed their volumes to create an overall volume measure. For cortical thickness, we averaged the right and left measurements for the entorhinal, fusiform, inferior temporal, middle temporal, inferior parietal, and precuneus regions and estimated the volume-weighted average of the six regional thicknesses to yield the cortical-thickness meta-ROI. To preserve temporal ordering between biological aging markers and neurodegeneration, all neuroimaging outcomes were derived from MRI scans obtained after baseline genomic data collection. Participants whose final MRI coincided with the genomic baseline visit were excluded. The average time between baseline genomic sampling and the last MRI visit was 4.26 ± 2.89 years.

### Biological Aging

#### Epigenetic Age

DNA methylation (DNAm) profiling was conducted on genomic DNA extracted from whole blood sample using the Illumina Infinium Human MethylationEPIC V1 BeadChip Array, following previously described protocols ^29^. Raw DNAm intensity data were normalized using the dasen method implemented within the‘wateRmelon’ R package. Quality control procedures were applied at both the probe and sample levels using the‘minfì and’DMRcatè R packages. CpG probes were excluded if they exhibited a detection p-value > 0.05 across all samples, were located within 5 base pairs of known single-nucleotide polymorphisms annotated with a minor allele frequency >0.01, were identified as cross-hybridizing probes, or were located on sex chromosomes (X or Y). Samples were excluded if their estimated bisulfite conversion efficiency fell below 85%. After applying these QC filters, 673,027 high-quality probes were retained for downstream analyses.

Epigenetic age was derived using CausAge clock, implemented via the‘Biolearn’ Python package ^4,37^. CausAge is derived from 586 CpG sites previously identified through Mendelian randomization analyses as having potential causal associations with the aging process ^4^. In all downstream statistical analyses, baseline chronological age was included as a covariate. Adjusting for chronological age in this framework is statistically equivalent to modeling epigenetic age acceleration, as it estimates the association between biological age and outcomes independent of chronological age.

#### Mitochondrial DNA copy number

Mitochondrial DNA copy number (mtDNAcn) quantification has been described in detail previously ^38^. Briefly, whole-genome sequencing CRAM files were processed using Mosdepth to estimate base coverage for both mtDNA and autosomal DNA. mtDNAcn was then calculated as twice the ratio of the mean mtDNA coverage to the mean autosomal coverage, thereby accounting for the diploid nature of the nuclear genome ([mean coverage of mtDNAcn / mean coverage of autosomes] x 2).

#### Telomere Length

Telomere length estimation and quality control have been previously described in detail ^30^. Briefly, genomic DNA was extracted from buffy coats or blood samples. Quantitative polymerase chain reaction (qPCR) was used to determine the relative telomere length as the ratio of telomere repeats to a single-copy gene (T/S ratio). The T/S ratios were adjusted for experimental variables, including plate, row, column, sample type (blood or buffy coat), and the storage time between DNA extraction and qPCR. The adjusted T/S ratios were then converted into base pairs using the formula: telomere base pairs = 3274 + 2413 × (T/S ratio).

#### Cell type Deconvolution

To account for cellular heterogeneity in blood, relative proportions for the six most abundant cell types found in blood (neutrophils, monocytes, NK cells, B cells, CD4+ T cells, and CD8+ T cells) were estimated from the bulk methylation data, using a reference-based deconvolution approach implemented via the‘Biolearn’ Python package. CpG sites for deconvolution were determined by identifying the 50 most distinctively hyper-and hypo-methylated sites per cell type (a total of 600 reference CpG sites), prioritized by the significance of their differential methylation.

#### Statistical Analysis

Differences in demographics, AD endophenotypes (cognitive function, CSF biomarkers, and neurodegeneration), and biological aging measures across diagnosis at study baseline were determined using a Kruskal-Wallis rank sum test, Pearson’s Chi-squared test, or Fisher’s exact test. Differences in biological aging markers across diagnostic groups were tested using one-way ANOVA with Tukey’s HSD post hoc tests. Pairwise Pearson correlation coefficients were computed between biological aging markers and AD endophenotypes using complete-case data. Statistical significance was determined using FDR-adjusted p-values with adjusted q < 0.05 considered significance.

For each AD endophenotype, we: 1) evaluated the independent effect of each biological aging marker in separate regression models; 2) assessed their joint effect by including all markers in a single regression model; and 3) examined interactions by evaluating two-way interactions (mtDNAcn × TL; mtDNAcn × CausAge; TL × DNAm age) and a three-way interaction (mtDNAcn × TL × CausAge). All regression and mixed-effects models were estimated using model-wise complete-case data, such that participants with missing values in any variable included in a given model were excluded from that model’s fit. Covariate data were largely complete, with minimal missingness for most variables (e.g., baseline CDR-SB missing in 10 participants in cognitive models). Missingness in other analyses primarily reflected availability of biomarker measurements (e.g., mtDNAcn, DNAm age, and telomere length) rather than incomplete covariate reporting. No imputation procedures were applied. Nominal significance was set at a = 0.05. Given the number of outcomes and models tested, multiple comparisons were controlled using Bonferroni correction within each outcome domain (cognitive, CSF, and MRI). For each domain, the number of tests (m) was defined as the total number of predictor–outcome pairs evaluated within that domain, including both main effects and interaction terms, and Bonferroni-adjusted p-values were computed accordingly (cognitive: m = 70; CSF: m = 21; MRI: m = 14).

Linear mixed-effects models were used to evaluate the association of TL, CausAge, and mtDNAcn with cognitive decline (global cognition and each cognitive domain) in participants with at least two cognitive visits. Both time (in years from baseline) and the intercept were included as fixed and random effects to assess the association of TL, mtDNAcn, and DNAm age and their interactions with baseline global cognitive function and cognitive decline. Models adjusted for baseline age x time, CDR-SB x time, *APOE* genotype (e2 carriers [e2/e2, e2/e3], e3/e3, and e4 carriers [e2/ e4, e3/e4, e4/e4]) x time, sex, education (high school, college, graduate level), and blood cell-type composition (monocytes, NK cells, B cells, CD4+ T cells, and CD8+ T cells; neutrophils were excluded to avoid multicollinearity). Baseline CDR-SB was included as a covariate to adjust for disease stage as it provides a finer gradation of clinical severity than simple clinical diagnosis or CDR global score, particularly in cognitively unimpaired and mild-impairment range ^39^. Including baseline chronological age as a covariate allows us to determine whether epigenetic age explains outcome variation independently of, and in addition to, chronological age. The linear mixed-effects formulas are available in supplementary methods.

Linear regression models were used to evaluate the cross-sectional associations of TL, mtDNAcn, and CausAge and their interactions with CSF Ab_42_, tau, and pTau_181_, adjusting for age, sex, CDR-SB, *APOE* genotype, and blood cell-type composition.

Linear regression models assessed the association of each biological aging marker and their interactions with cortical thickness and gray matter volume at the last visit, adjusting for age, sex, CDR-SB, months since baseline, *APOE* genotype, blood cell-type composition, and intracranial volume (for gray matter volume). MRI at last visit was used to strengthen the temporal ordering between exposure and outcome, with baseline biological-age markers measured first, and subsequent cortical-thickness or gray-matter volume reflects the cumulative neurodegenerative change that followed.

We conducted sensitivity analyses additionally adjusting for body mass index (BMI) and hypertension. We re-estimated all primary model specifications, including TL-only, DNAm-only, mtDNAcn-only, joint models, all two-way interactions, and the three-way interaction. Analyses were restricted to participants with non-missing BMI and hypertension (n = 629).

## Data availability

Data is available from https://ida.loni.usc.edu/.

Code is available at https://github.com/AndrewsLabUCSF/DNAm.

## Results

### Cohort Description

Biological aging measures for TL, mtDNAcn, and CausAge were available for 640 participants. The mean age of participants was 74.95 ± 7.62 years, with 44.2% female, 96.1% non-Hispanic White, 33.8% cognitively unimpaired, 51.6% diagnosed with MCI, 14.7% with dementia, and with longitudinal cognitive assessments for a duration of an average of 4.8 ± 3.1 years from the study baseline assessment. MRI data were available for 588 participants, while CSF biomarkers were available for 448 participants within one year of DNA isolation. TL did not differ across diagnostic groups. In contrast, significant differences in mtDNAcn and CausAge were observed; mtDNAcn was significantly lower in dementia participants compared to MCI, while CausAge was significantly higher in CU participants compared to MCI but lower than in dementia participants. AD endophenotypes were significantly worse in MCI and AD participants compared to CU participants.

Pairwise correlations were used to assess relationships among biomarkers of aging (Figure 1). mtDNAcn and TL were weakly negatively correlated with chronological age, whereas CausAge was strongly positively correlated. Similarly, mtDNAcn and TL were weakly positively correlated with each other, but both were weakly negatively correlated with CausAge. We similarly evaluated pairwise correlations between biomarkers of aging and AD outcomes. CausAge was weakly negatively correlated with gray matter volume, cortical thickness, and global cognition, while TL and mtDNAcn were not significantly correlated with AD outcomes. Chronological age exhibited weak negative correlations CSF Aβ_42_, gray matter volume, cortical thickness, and global cognition.

**Figure 1:**
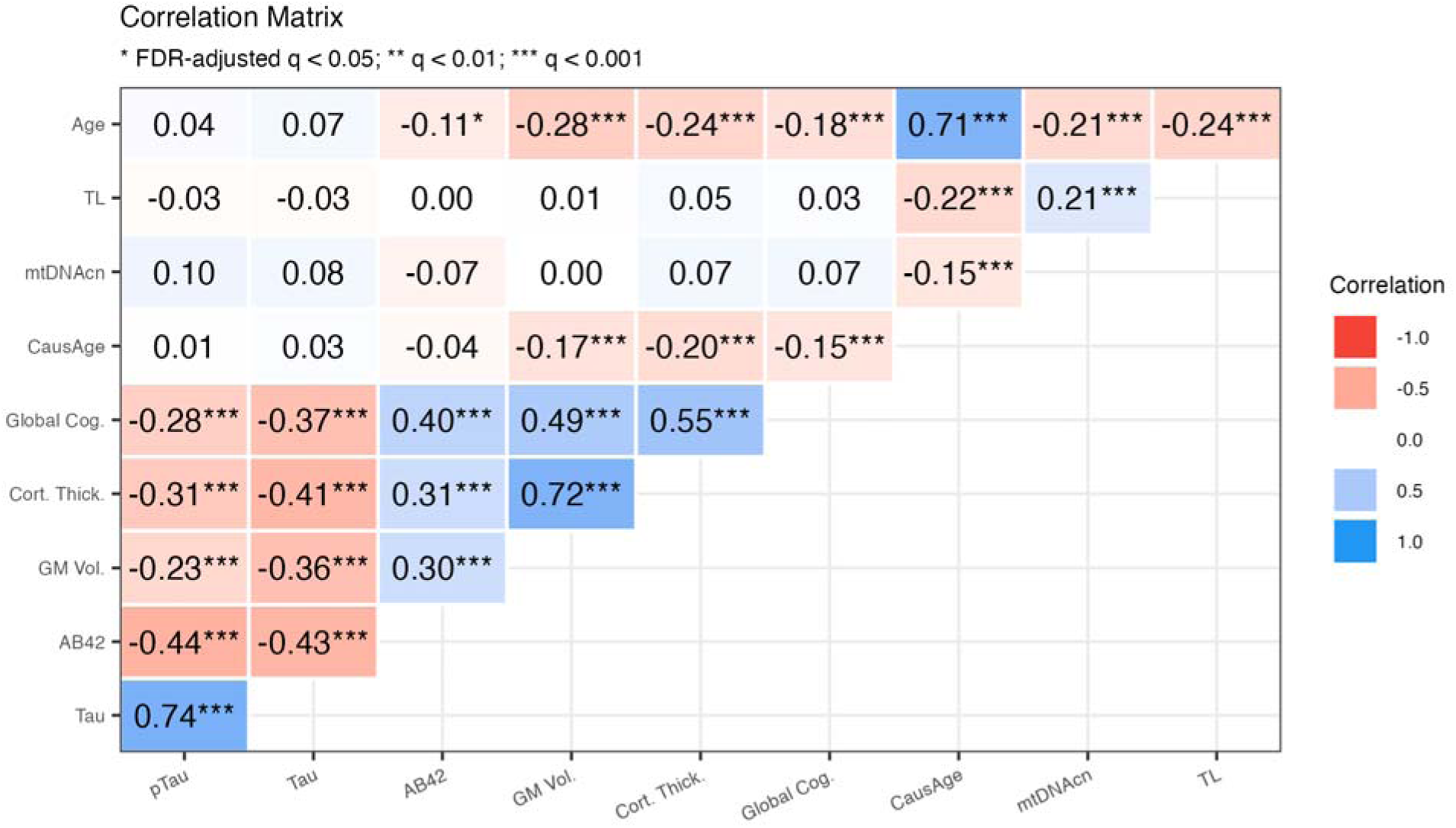
Pairwise correlations between biological aging markers and AD endophenotypes. Heatmap of Pearson correlation coefficients (r). Color intensity represents the magnitude and direction of associations. Statistical significance is denoted by asterisks based on FDR-adjusted p-values (*q < 0.05; **q < 0.01; ***q < 0.001). Effect sizes are categorized as very weak (|r| < 0.20), weak (0.20–0.39), moderate (0.40–0.59), and strong (≥0.60).

### TL and CausAge are not associated with AD endophenotypes; mtDNAcn is associated with tau pathology

Linear mixed-effects models were used to estimate the independent and joint associations of each biological aging marker with baseline cognitive function and its longitudinal change. No significant associations were observed for global cognitive function nor any specific cognitive domain (Figure 2; Table S1-5). Similarly, linear regression analyses were conducted to assess the cross-sectional associations of each marker, both independently and jointly, with CSF Aβ_42_, tau, and pTau_181_ (Figure 2; Table S6); higher mtDNAcn was associated with lower total tau and pTau_181_ [β =-0.24; p = 0.013*] levels. Finally, linear regression was used to evaluate the association of baseline TL, mtDNAcn, and CausAge with cortical thickness and gray matter volume at the last visit, and no significant associations were observed (Figure 2; Table S7). Results were consistent in sensitivity analyses, not adjusting for CDR-SB.

**Figure 2:**
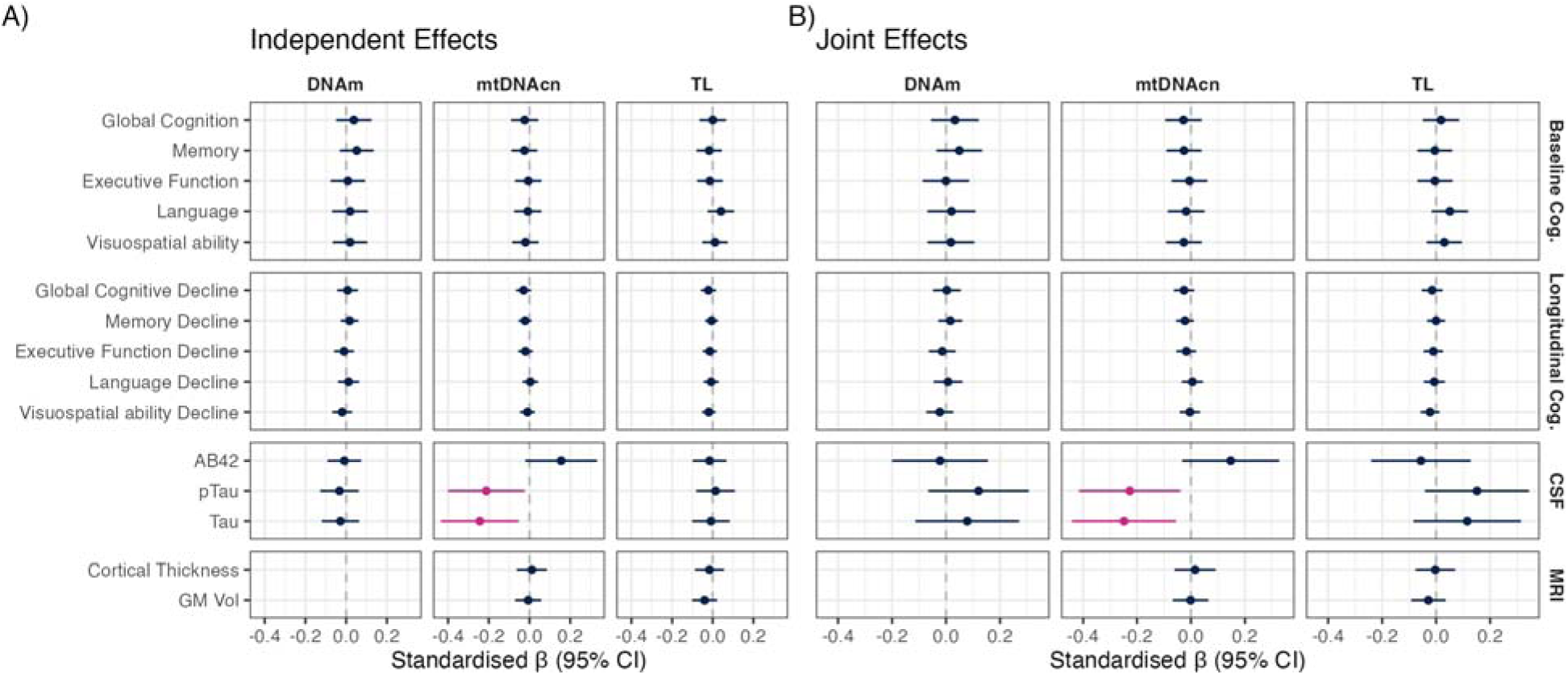
**Independent and joint association of biological aging markers with AD endophenotypes. *A) Independent Effects.*** Each bioage marker (DNAm, mtDNAcn, or TL) was entered in a separate model with the covariate set; β therefore reflects its stand-alone association with the outcome. ***B) Joint effects.*** All three markers were entered simultaneously; β estimates indicate the unique contribution of each marker after mutual adjustment. Magenta denotes statistically significant (p < 0.05) effects.

### mtDNAcn moderates the association of TL and CausAge with cognitive performance and gray matter volume

We evaluated the pairwise and three-way interactions between each biological aging marker and AD endophenotypes (Figure 3a). Significant interactions were observed between mtDNAcn and TL on baseline global cognition, memory, and executive function, such that, at higher mtDNAcn, shorter telomere length was associated with lower cognitive function (Figure 3b; Table S1-3). A significant interaction was also observed between mtDNAcn and DNAm age on baseline language ability; at higher mtDNAcn, for individuals with an older epigenetic age was associated with worse language performance (Figure 3c; Table S4). A significant three-way interaction was also observed, such that higher mtDNAcn was associated with reduced gray matter volume in the context of older DNAm age and longer TL Figure 3d; Table S7).

**Figure 3:**
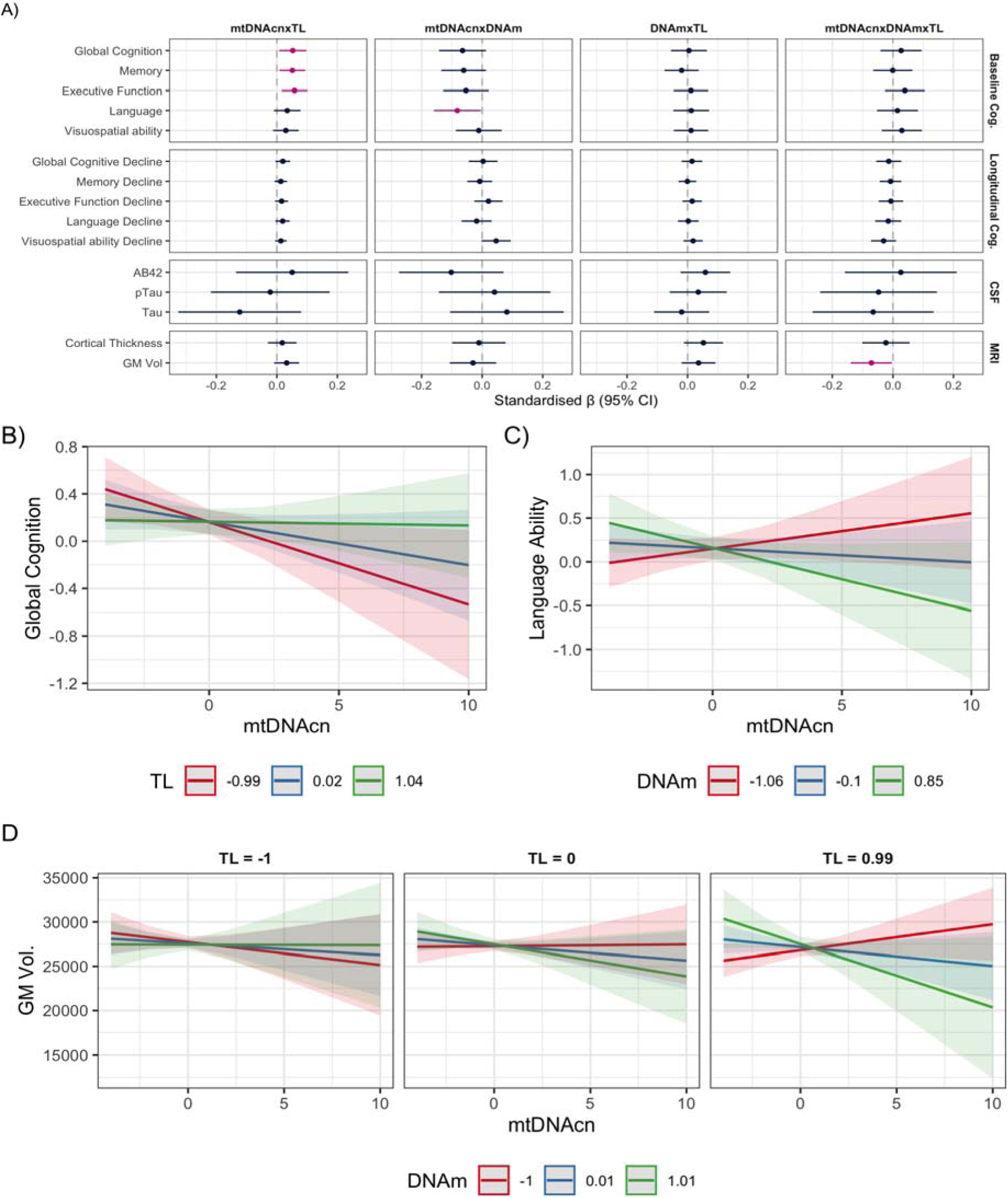
**Interactive effects among biological-age markers on AD endophenotypes: *A) Coefficient plot.*** Standardized regression coefficients (β, circles) with 95 % confidence intervals (bars) for every two-way interaction (mtDNAcn × TL, mtDNAcn × DNAm, DNAm × TL)and for the three-way interaction (mtDNAcn × DNAm × TL) across baseline cognition, longitudinal cognitive decline, CSF biomarkers, and MRI measures. Magenta denotes statistically significant interactions. ***B) mtDNAcn × TL on global cognition.*** Predicted baseline global-cognition z-score versus mtDNAcn, with telomere length held at –1 SD (red, “short”), 0 SD (blue, “average”), or +1 SD (green, “long”). ***C) mtDNAcn × DNAm on language ability.*** Predicted baseline language-ability z-score versus mtDNAcn, with DNAm-age acceleration fixed at –1 SD (red, “younger”), 0 SD (blue), or +1 SD (green, “older”). ***D) Three-way interaction on grey-matter volume.*** Each facet fixes telomere length at –1, 0, or +1 SD (left-to-right). Within each facet, lines depict the mtDNAcn–grey-matter-volume slope at DNAm–1 SD (red), 0 SD (blue), and +1 SD (green).

## Discussion

Markers of biological aging are not isolated indicators of organismal aging but rather interconnected components of complex processes that drive organismal health. Here, we sought to comprehensively evaluate the joint and interactive effects of TL, mtDNAcn, and DNAm age on AD endophenotypes. We observed that TL, and DNAm age were not associated with cognitive decline, CSF Aβ_42_, tau, and pTau_181_, or meta-ROI for cortical thickness and gray matter volume, while higher mtDNAcn was associated with lower tau and pTau_181_. We also observed significant interactions, with mtDNAcn moderating the effect of TL and DNAm on cognitive function and gray matter volume.

We observed that chronological age and CSF biomarkers of AD pathology showed the strongest correlations with cognitive decline and brain morphometry, whereas biomarkers of biological aging were either not correlated or, in the case of epigenetic age, only weakly correlated. This pattern suggests that each biological aging biomarker captures only one dimension of aging rather than overall aging burden, and that these markers may be more informative in combination or through interaction effects than as strong standalone correlates of AD endophenotypes.

Prior studies that have used ADNI to investigate the association of TL ^30,40,41^, DNAm age ^5,6,42–45^, and mtDNAcn ^38^ on AD endophenotypes have produced mixed results. TL was not associated with baseline MCI/AD, episodic memory performance, CSF Ab and tau, nor with regional gray matter volume and cortical thickness ^30,40^. However, longer telomere length was associated with faster decline in executive function and interacted synergistically with CSF Ab, CSF tau, and *APOE* ε4 to worsen executive function ^40^. In contrast, shorter telomere length was associated with older predicted brain age but not cognitive age ^41^.

Only one ADNI study has examined mtDNAcn, with higher mtDNAcn being associated with lower CSF Aβ_42_ but not with FDG-or Amyloid-PET, or A/T/N fluid biomarker ^38^. Stratified analyses suggested mtDNAcn was associated with FDGLPET, amyloid-PET, and CSF t-Tau and pTau_181_ in women but not in men. In contrast, we did not observe significant mtDNAcn-CSF Aβ_42_ associations after covariate adjustment. This discrepancy likely reflects differences in modeling and sampling. Specifically, our CSF models adjusted for baseline clinical severity and blood cell-type composition, and we restricted CSF measures to those obtained within one year of genomic sampling to improve temporal alignment, which reduced sample size and power. In addition, our primary analyses emphasized multivariable and interaction models that simultaneously accounted for TL and DNAm age rather than single-marker or stratified models. Together, these differences suggest mtDNAcn–Aβ_42_ associations in ADNI may be sensitive to model specification.

DNAm age findings vary with the clock and with the definition of ageLacceleration (absolute vs relative vs intrinsic). Third-generation clocks such as DunedinPACE are more consistently associated with increased risk of MCI/AD and worse cognitive performance, whereas first-(Horvath and Hannum) and second-generation clocks (PhenoAge, GrimAge) generally show weak or domainLspecific effects ^5,43^. In contrast, PhenoAge was associated with incident diagnosis and faster rates of global cognitive decline, while GrimAge was predominantly associated with cortical thinning and WMH burden ^45^. Few studies have evaluated the association of epigenetic clocks with CSF A/T/N biomarkers ^44^. Additionally, CausAge has not previously been applied in ADNI. Unlike first-and second-generation clocks trained on chronological age or mortality, CausAge is enriched for CpG sites with putative causal relevance to aging processes. As such, it may capture mechanistic aging biology rather than cumulative disease burden, potentially explaining why DNAm age did not emerge as an independent predictor in our models but instead modified mtDNAcn associations in interaction analyses. Collectively, heterogeneity in quantifying epigenic age and modelling choices appears to underlie the inconsistent associations observed across studies.

For the majority of AD endophenotypes, we did not identify any associations with biological aging markers; however, we did observe several interactions with mtDNAcn. Significant interaction-only models can arise when the effect of a predictor changes direction or magnitude across levels of the moderator, such that the positive and negative components cancel when averaged, even though the context-specific effects are strong. First, we observed that among participants with shorter telomere length, higher mtDNAcn was associated with lower cognitive function. Similarly, in participants with an older epigenetic age, higher mtDNAcn was associated with worse language ability. Finally, higher mtDNAcn was associated with reduced gray matter volume in participants with longer telomeres and older epigenetic age.

A plausible mechanism is a compensatory increase in mtDNA replication that is initiated when telomere attrition activates p53, suppresses PGC-1α (the master driver of mitochondrial biogenesis), and down-regulates TFAM, thereby compromising oxidative phosphorylation ^20,21^. Early in this cascade, a transient drop in TFAM-to-mtDNA ratio results in less compact mitochondrial nucleoids, which facilitates additional mtDNA replication and transcription to restore ETC protein levels and ATP output ^46^. Consequently, the observed interaction between shorter telomere attrition and increased mtDNAcn may reflect stress-induced compensation to restore mitochondrial function, rather than robust mitochondrial health ^47^. When this compensatory response becomes chronic, elevated mtDNAcn may reflect persistent bioenergetic stress rather than resilience ^15,47^. In neuronal tissue, disrupted bioenergetics can limit ATP availability, impairing synaptic function and neurotransmitter synthesis, release, and uptake ^47^. Overtime, this energy constraint can reduce neuronal maintenance capacity and increase vulnerability to neurodegenerative processes, which may manifest as gray matter volume loss ^33,47^. These structural changes can then contribute to declines in cognitive function, particularly in global cognitive domains ^17,40,47^. Overall, the observed interaction patterns support a context-dependent model in which elevated mtDNAcn marks metabolic stress in biologically aged systems, with downstream effects on both structural brain integrity and cognitive performance.

Our findings should be interpreted alongside some study limitations. First, the number of ADNI participants with available genomics to estimate TL, DNAm age, and mtDNAcn is limited, resulting in a small sample size and limited power to detect significant effects. Due to limited sample size for longitudinal mixed-effects models, we were unable to perform sex-stratified analyses, and future studies in larger cohorts will be necessary to determine whether the observed biomarker associations differ between men and women. In addition, CSF biomarker availability in the ADSP-PHC release used for this study was limited to Aβ_42_, total tau, and p-Tau_181_. More sensitive biomarkers of AD pathology (e.g. Aβ_42_/Aβ_40_ and p-Tau_217_) may improve detection of associations between aging biomarkers and amyloid or tau pathology should be evaluated in future studies. Second, chronological age is strongly correlated with clinical diagnosis and disease stage in ADNI, with MCI participants being younger on average than cognitively normal individuals. Although models adjusted for baseline age and clinical severity, some observed associations may partly reflect age-related differences in disease stage rather than age-independent effects, particularly given differences in age distributions across diagnostic groups. Age-stratified and nonlinear age sensitivity analyses were not feasible due to the small sample size and should be evaluated in larger cohorts. Third, our measures of biological aging were derived from DNA extracted from whole blood and not brain tissue. As such, we assume that blood-derived telomere attrition, epigenetic alterations, and mitochondrial dysfunction reflect systemic biological aging ^48,49^. As such, we are unable to determine if interactions between biological aging measures in brain tissue may influence AD pathogenesis. Fourth, residual confounding by exposome risk factors (e.g., socioeconomic context, health behaviors, and comorbidities) is possible, but ADNI has limited availability of these variables. To partially address this concern, we conducted additional sensitivity analyses adjusting for BMI and hypertension as representative cardiometabolic risk factors, and the overall pattern of results remained consistent. Additional adjustment could attenuate some associations; however, expanding covariate sets in observational analyses can also introduce overadjustment or collider bias when variables lie on the causal pathway or are influenced by disease progression. Replication in cohorts with more detailed exposome measures, ideally alongside causal modeling frameworks, will be needed to evaluate robustness to alternative adjustment sets and to better distinguish confounding from mediation. Finally, participants in ADNI are primarily non-Hispanic White and have a higher education than the general population. As such, our findings require further replication in diverse prospective population-based cohorts.

Despite these limitations, this study exhibits multiple strengths, including the breadth of genomic, cognitive, imaging, and fluid biomarker data available, which allowed us to conduct comprehensive analyses of the joint, independent, and interactive effects of TL, DNAm age, and mtDNAcn on AD pathogenesis.

In conclusion, this study examined how three genomic hallmarks of biological aging - telomere length, DNAm age, and mtDNAcn – influence AD pathogenesis. Independently, only mtDNAcn was associated with tau pathology; however, we also found that DNAm age and TL moderated the effect of mtDNAcn on cognitive function and gray matter volume. These findings argue for a systems-level view of brain aging. The classical “primary hallmarks” of aging—genomic instability, epigenetic alteration, and impaired proteostasis—do not harm neurons or glial cells in isolation; rather, they interact, triggering secondary cascades such as mitochondrial dysfunction, disrupted inter-cellular signaling, and chronic inflammation. Future work should therefore model multiple aging pathways simultaneously and test how their convergence accelerates—or, if targeted therapeutically, slows—AD pathogenesis.

## Acknowledgments

ADNI

*Data used in preparation of this article were obtained from the Alzheimer’s Disease Neuroimaging Initiative (ADNI) database (adni.loni.usc.edu). As such, the investigators within the ADNI contributed to the design and implementation of ADNI and/or provided data but did not participate in analysis or writing of this report. A complete listing of ADNI investigators can Be found at: http://adni.loni.usc.edu/wp-content/uploads/how_to_apply/ADNI_Acknowledgement_List.pdf

Data collection and sharing for this project was funded by the Alzheimer’s Disease Neuroimaging Initiative (ADNI) (National Institutes of Health Grant U01 AG024904) and DOD ADNI (Department of Defense award number W81XWH-12-2-0012). ADNI is funded by the National Institute on Aging, the National Institute of Biomedical Imaging and Bioengineering, and through generous contributions from the following: AbbVie, Alzheimer’s Association; Alzheimer’s Drug Discovery Foundation; Araclon Biotech; BioClinica, Inc.; Biogen; Bristol-Myers Squibb Company; CereSpir, Inc.; Cogstate; Eisai Inc.; Elan Pharmaceuticals, Inc.; Eli Lilly and Company; EuroImmun; F. Hoffmann-La Roche Ltd and its affiliated company Genentech, Inc.; Fujirebio; GE Healthcare; IXICO Ltd.;Janssen Alzheimer Immunotherapy Research & Development, LLC.; Johnson & Johnson Pharmaceutical Research & Development LLC.; Lumosity; Lundbeck; Merck & Co., Inc.;Meso Scale Diagnostics, LLC.; NeuroRx Research; Neurotrack Technologies; Novartis Pharmaceuticals Corporation; Pfizer Inc.; Piramal Imaging; Servier; Takeda Pharmaceutical Company; and Transition Therapeutics. The Canadian Institutes of Health Research is providing funds to support ADNI clinical sites in Canada. Private sector contributions are facilitated by the Foundation for the National Institutes of Health (www.fnih.org). The grantee organization is the Northern California Institute for Research and Education, and the study is coordinated by the Alzheimer’s Therapeutic Research Institute at the University of Southern California. ADNI data are disseminated by the Laboratory for Neuro Imaging at the University of Southern California.

## Author Contributions

SJA: Design or conceptualization of the study; analysis or interpretation of the data; drafting of the manuscript for intellectual content. BAM: major role in the acquisition of data; revising the manuscript for intellectual content. TT: major role in the acquisition of data; revising the manuscript for intellectual content. LWB: analysis or interpretation of the data; revising the manuscript for intellectual content. AER: Design or conceptualization of the study; revising the manuscript for intellectual content. XZ: major role in the acquisition of data; revising the manuscript for intellectual content. MS: major role in the acquisition of data; revising the manuscript for intellectual content. DT: analysis or interpretation of the data; revising the manuscript for intellectual content. KY: analysis or interpretation of the data; revising the manuscript for intellectual content

