## Supplementary Methods for "Integrative effects of Telomere Length, Epigenetic Age, and Mitochondrial DNA abundance in Alzheimer’s Disease"

**Linear Mixed Effects Models**

Longitudinal cognitive trajectories were modeled using linear mixed-effects models with random intercepts and random slopes for time at the participant level.

Let $Y_{ij}$ denote the cognitive outcome for participant $i$ at visit$j$. The general model can be written as:

$$Y_{ij}=X_{ij}\beta+Z_{ij}b_{i}+\epsilon_{ij}$$

Where $Y_{ij}$ is the outcome variable; $X_{ij}$ is the fixed-effects design matrix; $\beta$ is the vector of fixed-effect coefficients; $Z_{ij}$ is the random-effects design matrix; $b_{i}\sim N\left( 0, D \right)$ is the vector of participant-specific random effects;$\epsilon_{ij}\sim N\left( 0, \sigma^{2} \right)$ is the residual error; $Cov \left( b_{i}, \epsilon_{ij} \right)$ = 0.

Random effects included a participant-specific intercept and slope for years since baseline.

Time was defined as centered years since baseline $\left( yrs_{-}c \right)$. For longitudinal cognition, fixed-effects covariates included: $\left( yrs_{-}c \right)$; baseline age $\left( z_{-}age_{-}bl \right)$ and its interaction with time ($\left( z_{-}age_{-}bl \times yrs_{-}c \right)$; sex; education category $\left( educ_{-}cat \right)$; baseline clinical severity $\left( cdrsb_{-}bl \right)$ and its interaction with time $\left( cdrsb_{-}bl \times yrs_{-}c \right)$); APOE genotype $\left( apoe \right)$ and its interaction with time $\left( apoe \times yrs_{-}c \right)$; and baseline immune cell-type proportions $\left( nk_{-}cell_{-}bl, b_{-}cell_{-}bl, cd4_{-}t_{-}cell_{-}bl, cd8_{-}t_{-}cell_{-}bl, monocyte_{-}bl \right)$.

All biological aging markers were standardized (z-scored) at baseline:

Telomere length $\left( z_{-}tl_{-}bl \right)$, Epigenetic age (CausAge; $z_{-}dnam_{-}bl$), Mitochondrial DNA copy number ($z_{-}mtcn_{-}bl$)

The linear mixed effects model equation was as follows:

**Model 1 (independent)**

$$Y_{ij} = \beta_{0}+ \beta_{1}\left( yrs_{-}c_{ij} \right) + \beta_{2}\text{M}_{\text{i}} + \beta_{3}\left( \text{M}_{\text{i}} \times yrs_{-}c_{ij} \right) + \text{γ}^{\text{T}}\text{C}_{\text{ij}} + b_{0i} + b_{1i}\left( yrs_{-}c_{ij} \right) + \epsilon_{ij}$$

Equivalent models were run separately for TL, DNAm/ CausAge and mtDNAcn.

**Model 2 (joint)**

The joint model included all three biological aging markers and their interactions with time:

$$Y_{ij} = \beta_{0} + \beta_{1}\left( yrs_{-}c_{ij} \right) + \beta_{2}\left( z_{-}tl_{-}bl_{i} \right) + \beta_{3}\left( z_{-}dnam_{-}bl_{i} \right) + \beta_{4}\left( z_{-}mtcn_{-}bl_{i} \right) + \beta_{5}\left( z_{-}tl_{-}bl_{i} \times yrs_{-}c_{ij} \right) +$$

$$\beta_{6}\left( z_{-}dnam_{-}bl_{i} \times yrs_{-}c_{ij} \right) + \beta_{7}\left( z_{-}mtcn_{-}bl_{i} \times yrs_{-}c_{ij} \right) + \gamma^{T}C_{ij} + b_{0i}+b_{1i}\left( yrs_{-}c_{ij} \right)+\epsilon_{ij}$$

**Model 3 (2-way interaction)**

To evaluate pairwise interactions among biological aging markers, we fit three separate models, each including a pairwise interaction term and its interaction with time. For a generic pair $\left( A_{i}, B_{i} \right)$, where $A_{i}$, $B_{i} \in$ $\left\{ z_{-}tl_{-}bl_{i}, z_{-}dnam_{-}bl_{i}, z_{-}mtcn_{-}bl_{i} \right\}$, the model was specified as:

$$Y_{ij} = \beta_{0} + \beta_{1}\left( yrs_{-}c_{ij} \right) + \beta_{2}A_{i} + \beta_{3}B_{i} + \beta_{4}\left( A_{i}\times B_{i} \right)$$

$$+ \beta_{5}\left( A_{i} \times yrs_{-}c_{ij} \right) + \beta_{6}\left( B_{i} \times yrs_{-}c_{ij} \right) + \beta_{7}\left( A_{i} \times B_{i} \times yrs_{-}c_{ij} \right)$$

$$+ \gamma^{T}C_{ij} + b_{0i} + b_{1i}\left( yrs_{-}c_{ij} \right) + \epsilon_{ij}$$

Separate models were estimated for the following marker pairs:

- TL × DNAm
- TL × mtDNAcn
- DNAm × mtDNAcn

Each model therefore evaluated whether the interaction between two biological aging markers modified baseline cognition and longitudinal cognitive change.

**Model 4 (3-way interaction)**

To test whether the combined interaction among all three biological aging markers influenced cognitive trajectories, we fit a model including the three-way interaction and its interaction with time:

$$Y_{ij}=\beta_{0} + \beta_{1}\left( yrs_{-}c_{ij} \right) + \beta_{2}\left( z_{-}tl_{-}bl_{i} \right) + \beta_{3}\left( z_{-}dnam_{-}bl_{i} \right) + \beta_{4}\left( z_{-}mtcn_{-}bl_{i} \right)$$

$$+ \beta_{5}\left( z_{-}tl_{-}bl_{i} \times z_{-}dnam_{-}bl_{i} \right) + \beta_{6}\left( z_{-}tl_{-}bl_{i} \times z_{-}mtcn_{-}bl_{i} \right) + \beta_{7}\left( z_{-}dnam_{-}bl_{i} \times z_{-}mtcn_{-}bl_{i} \right)$$

$$+ \beta_{8}\left( z_{-}tl_{-}bl_{i} \times z_{-}dnam_{-}bl_{i} \times z_{-}mtcn_{-}bl_{i} \right)$$

$$+ \beta_{9}\left( z_{-}tl_{-}bl_{i} \times yrs_{-}c_{ij} \right) + \beta_{10}\left( z_{-}dnam_{-}bl_{i} \times yrs_{-}c_{ij} \right) + \beta_{11}\left( z_{-}mtcn_{-}bl_{i} \times yrs_{-}c_{ij} \right)$$

$$+ \beta_{12}\left( z_{-}tl_{-}bl_{i} \times z_{-}dnam_{-}bl_{i} \times yrs_{-}c_{ij} \right) + \beta_{13}\left( z_{-}tl_{-}bl_{i} \times z_{-}mtcn_{-}bl_{i} \times yrs_{-}c_{ij} \right)$$

$$+ \beta_{14}\left( z_{-}dnam_{-}bl_{i} \times z_{-}mtcn_{-}bl_{i} \times yrs_{-}c_{ij} \right) + \gamma^{T}C_{ij} + b_{0i} + b_{1i}\left( yrs_{-}c_{ij} \right) + \epsilon_{ij}$$

All lower-order terms were retained when higher-order interaction terms were included, consistent with hierarchical model specification.
